# Forecasting patient-specific tumor response using patient-reported outcomes in non-small cell lung cancer

**DOI:** 10.64898/2026.01.29.26345069

**Authors:** Diya J. Upadhyaya, Matthew B. Schabath, Aasha I. Hoogland, Renee Brady-Nicholls

## Abstract

**Purpose:** Patient-reported outcomes (PROs) provide a quantitative measure of a patient’s quality of life, directly from the patient without external influence or interpretation. Prior studies have demonstrated correlations between individual PROs and cancer treatment response. However, this area of research is still highly understudied, and patient data often goes ignored. Our previous work has shown how changes in insomnia can be used to make binary decisions about a patient’s future volume response. Here, we expand upon that work to determine precisely when treatment progression will occur, providing an opportunity for clinicians to intervene sooner.

**Experimental Design:** This study analyzed PROs and tumor volume data collected from 80 NSCLC patients undergoing immunotherapy to determine how PRO dynamics could inform when volumetric treatment progression would occur. We calibrated the tumor growth inhibition (TGI) model to patient-specific tumor volume dynamics for all volume measurements using a leave-one-out cross-validation approach. Growth parameters were divided based on progression status and sampled depending on changes in patient-reported insomnia. A cutoff analysis was performed to determine the optimal cutoff for distinguishing between responders and non-responders. Predictions were made for the *N*^*th*^ patient and categorized using the cutoff.

**Results:** This study demonstrated that incorporating patient-specific changes in insomnia with a mathematical model of volume changes can predict patient response with a 72.2% true positive rate and 71.3% overall accuracy, on average 6-8 weeks sooner.

**Conclusion:** Using this innovative framework, we can predict precisely when progression occurs, giving clinicians the opportunity to intervene beforehand.

## Introduction

Lung cancer is the second most common cancer in the U.S., after breast and prostate cancers, but remains the leading cause of cancer death, with more than half of lung cancer patients dying within a year of their diagnosis [5]. The five-year survival rate remains less than 20% [5]. Non-small cell lung cancer accounts for more than 80% of lung cancer diagnoses [4]. At least 40% of NSCLC patients are stage IV at the time of diagnosis; consequently, treatment typically aims to improve survival and reduce cancer-associated adverse events [10]. Accordingly, biomarkers are essential for assessing a patient’s response to treatment longitudinally.

Biomarkers are frequently divided into diagnostic, prognostic, and predictive depending on their ultimate use. Due to tumor heterogeneity and irreproducible results, prognostic molecular biomarkers in non-small cell lung cancer are often left out of routine use in treatment [8]. Therefore, non-molecular prognostic biomarkers need to be evaluated. Patient-reported outcomes (PROs) assess a patient’s quality of life through directly reported symptomology questionnaires or surveys. These measures allow patients to report their symptoms without the interference of physicians or any other party, providing the opportunity to play a more direct role in their treatment. PROs serve as a noninvasive prognostic biomarker in several cancers. Previous research indicates its high correlation with volume dynamics in non-small cell lung cancer [3, 6, 2, 1]. In our previous study, we demonstrated how changes in insomnia could be used to predict whether or not a patient would progress at their next volume scan with 77% accuracy, on average, 45 days before the next scan [3].

Although insomnia has shown potential as a predictor of stable disease, its capacity to predict resistance is less promising, with only a 28% accuracy. It also neglects a timeline as to when progression will occur. To remedy this, we propose integrating tumor volume dynamics, using a mathematical model, with PRO dynamics to forecast tumor dynamics over time. We hypothesize that by incorporating insomnia changes into a mathematical model of tumor volume dynamics, we will provide an actionable and accurate biomarker of patient response.

## Materials and Methods

### Clinical Data and Study Design

This study included data from NSCLC patients undergoing immune checkpoint inhibitor therapy at Moffitt Cancer Center. Patients completed PRO surveys biweekly, assessing 27 symptoms associated with immunotherapy and cancer progression using a 5-point Likert scale with a score of 0 indicating no symptom severity and a score of 4 indicating severe symptomology. This study was conducted in accordance with the U.S. Common Rule and the Declaration of Helsinki, and the study protocol was approved by the Institutional Review Board (Advarra, Inc).

Approximately every month, patients underwent computed tomography (CT) scans to measure tumor volume, which evaluated using the Immunotherapy Response Evaluation Criteria in Solid Tumors (iRECIST) criteria. According to iRECIST criteria, progressive disease is defined by a 20% increase from the nadir, or lowest volume point, of the sum of the longest diameters in tumors, which translates to a 72.8% increase in tumor volume. Progressive disease can be further broken down into unconfirmed and confirmed progressive disease by a sequential increase in tumor volume at the next CT scan [9]. Further information is provided in the paper by Bhatt et al. [3].

The clinical data used to conduct this study are available upon reasonable request to the corresponding author (RBN).

### Mathematical Model

Following Glazar et al., we utilized a tumor growth inhibition (TGI) model to describe the effects of therapy on tumor volume [**7**]. This model has been previously used in several cancers, including NSCLC [7]. The model describes exponential growth of volume, decay due to treatment, and an eventual loss of inhibition rate over time due to an evolution of resistance. This model is as follows:

**Tumor burden (Volume):** 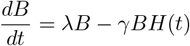

**Treatment Sensitivity:** 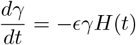

**Treatment:** 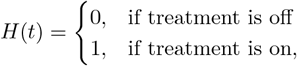

with initial conditions *B*_0_ and *γ*_0_. Treatment is modeling using the step function *H*(*t*).

### Model Calibration

We used the Levenberg-Marquardt, executed in MATLAB, to determine model parameters that minimized the least squares error between model simulation and patient data. The model was fit over all volume points for all 80 patients. As the aim of our study is to make patientspecific response predictions, we first needed to minimize the degrees of freedom. To do so, we used a nested optimization to determine which parameters should vary between patients and which should be uniform across all time points. The Akaike Information Criteria (AIC) was used to compare the model fits, given the number of degrees of freedom.

### Model Prediction

#### Without PROs

To generate predictions of treatment response, we utilized a leave-one-out (LOO) approach.

Following the pipeline in Figure 3, the LOO approach was used to test the robustness of the model. At each iteration of the code, a single patient was left out as the testing set and then analyzed against the training set, i.e., the rest of the patients. The analysis considered each patient from one volume measurement to the subsequent measurement. Therefore, predictions were divided into the pretreatment measurement to the first on-treatment volume measurement and from the pretreatment measurement to the second on-treatment volume measurement. As only three patients progressed from the third to the fourth measurement, they were excluded due to the imbalanced data. From volume measurement one to two, a total of 80 patients underwent the leave-one-out approach, generating 80 predictions.

Using this LOO approach, *N* − 1 patients were optimized over for all volume points. *λ* values were stored in a CDF curve while *γ*_0_ and *ϵ* were uniformly fixed. The CDF curve was randomly sampled 100 times to generate 100 prediction curves. Any curve with a 72.8 % increase from the nadir was considered a progressive curve, and then the total number of curves exhibiting resistance divided by the 100 curves was deemed the resistance rate.

To predict up to the third volume measurement, we repeated the same steps after removing the patients who only had two data points or who had already progressed from one to two.

After recording the resistance rates from each prediction for patients from the first to the second volume measurement, a cutoff analysis was performed to determine a threshold for progressive disease on the training set. A threshold for each iteration of the LOO was determined by maximizing the Youden Index to maximize the specificity and sensitivity in each dataset. Each cutoff value was then tested on the left-out patient to classify it as resistant or responsive. The predictions were then classified as true positive, false positive, true negative, or false negative and recorded.

#### With PROs

In our previous study [3], we found that increases in insomnia between volume measurements were indicative of progressive disease, while decreasing or stable levels of insomnia were predictive of response. We decided to build upon this and incorporate insomnia changes into the TGI model to predict patient-specific response dynamics.

We utilized the model parameters from the LOO optimization to store in the CDF curves. The CDF plot consisted of two curves, as shown in Figure 3, where the black curve stores the *λ* values of responsive patients, while the red curve does so for resistant patients. The figure demonstrates a discernible difference between *λ* values for resistant and responsive patients with a pvalue of 4.7746e-05. However, because several responsive and resistant patients showed no change in insomnia at each volume point, the estimated resistance rates were disproportionately weighed down. To remedy this, we introduced a third CDF curve, specifically for patients with no insomnia change, that combined the *λ* values for responsive and resistant patients.

The present study aims to incorporate these seemingly disparate components, so a condition was added to the model to account for changes in insomnia. The pretreatment growth rate in patients who demonstrated an increase in insomnia severity was generated by randomly sampling from the red curve in the CDF, while those with a decrease in severity were sampled from the black curve. As previously mentioned, patients with no change in severity were sampled from a curve combining the pretreatment growth rate values of the two curves. Following Bhatt et al., each different change was counted as a different prediction; however, an increase would indicate progression, and no subsequent changes would be considered. Therefore, patients with different changes, such as a decrease in insomnia and then a subsequent increase, would have multiple predictions. We then followed the same process as detailed above regarding sampling the appropriate CDF curve 100 times, generating prediction curves, and performing a cut-off analysis.

## Results

### Model Accurately Describes Patient-Specific Response

The AIC values are shown in Table 1. The AICs were very similar when allowing for one patient-specific parameter. In line with Glazar et al., we allowed *λ* to be patient-specific, while *γ*_0_ and *ϵ* were set to be uniform across patients. After running a nested optimization function, we found the calibrated model parameters. Table 2 describes our model conditions.

**Table 1.** AIC values of model performance when choosing patient-specific parameters.

| Patient-Specific Parameter | AIC Values |
| --- | --- |
| $\lambda$ | 15.5536 |
| $\epsilon$ | 15.9130 |
| $\gamma_0$ | 15.6600 |

**Table 2.** Table of Tumor Growth Inhibition Model Parameters.

| Parameter | Uniform or Patient-Specific | Calibrated-Value | Description | Units |
| --- | --- | --- | --- | --- |
| $\lambda$ | Patient-Specific | $[0.005 \log(2)]$ | Pre-Treatment Growth Rate | $day^{-1}$ |
| $\gamma$ | Uniform | 0.0286 | Initial Treatment Sensitivity | $day^{-1}$ |
| $\epsilon$ | Uniform | 0.0052 | Rate of Evolution of Resistance | $day^{-1}$ |
| $B_0$ | Patient-Specific | $[0.2761 \ 176.4110]$ | Initial Tumor Burden (Volume) | $cm^3$ |

Figure 1A illustrates that the model effectively captures the dynamics across patients. Patients 1 and 48 are resistant patients who progress at the third volume point and, according to the iRECIST criteria, have confirmed progression at the fourth volume point. Patients 18 and 91 are both responsive patients with varying amounts of volume data. After fitting for all patients, we divided the cohort into responsive and resistant patients, and Figure 1B shows how higher pre-treatment growth rates are found in resistant patients.

**Figure 1:**
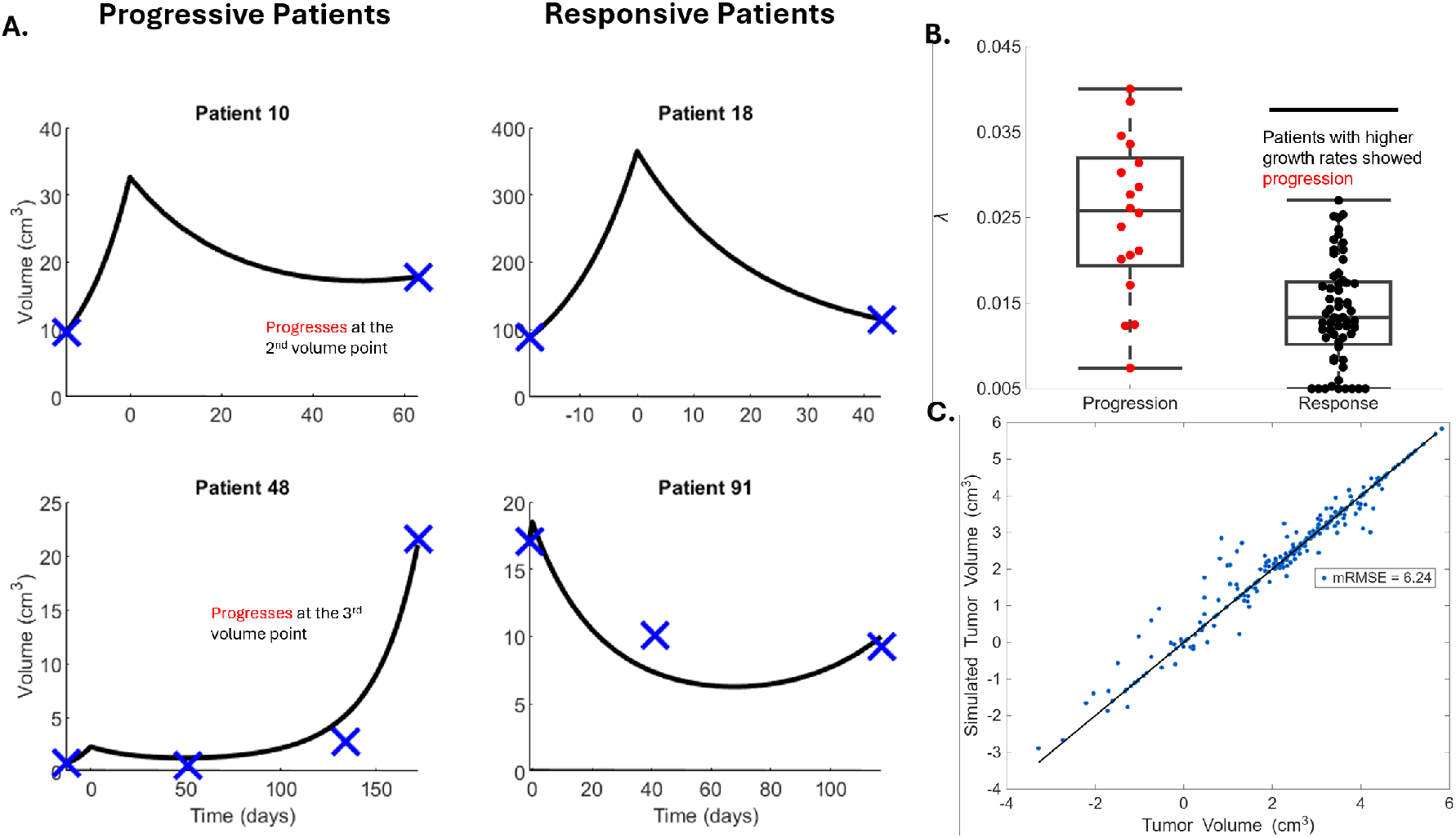
Model calibration results. **A**. Model fits to two progressive patients (left) and two responsive patients (right). **B**. Patient-specific growth rates are significantly different between patients who progress and those who respond. **C**. Actual versus estimated volumes for all patients.

### TGI Model Alone Cannot Predict Treatment Response

Figure 2 shows the receiver operating characteristic curve of the prediction simulations generated with no PROs. The plot is able to illustrate the diagnostic ability of the model alone by mapping the sensitivity (true-positive rate) against 1− specificity (false-positive rate).

**Figure 2:**
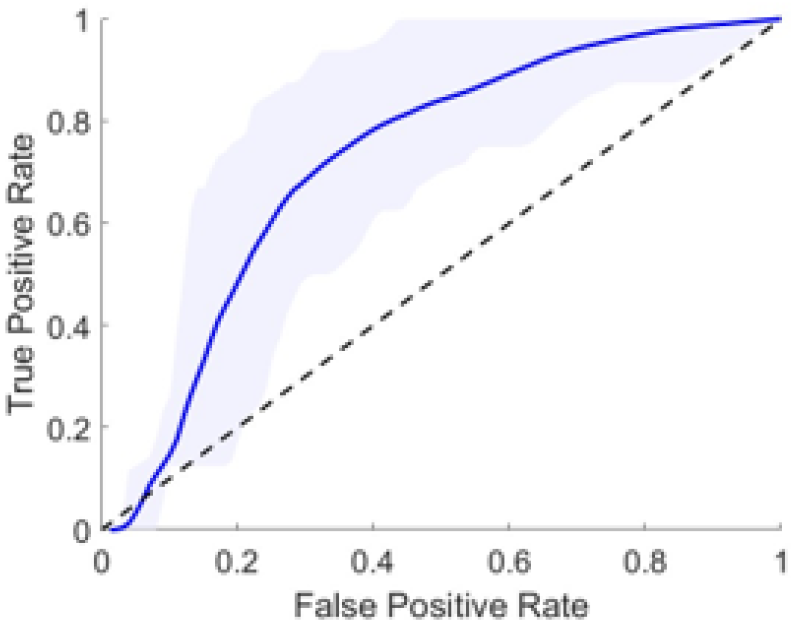
ROC curve of predictions using TGI model. The TGI model itself was able to increase sensitivity to 75% however, specificity dropped to 65.3%.

**Figure 3.**
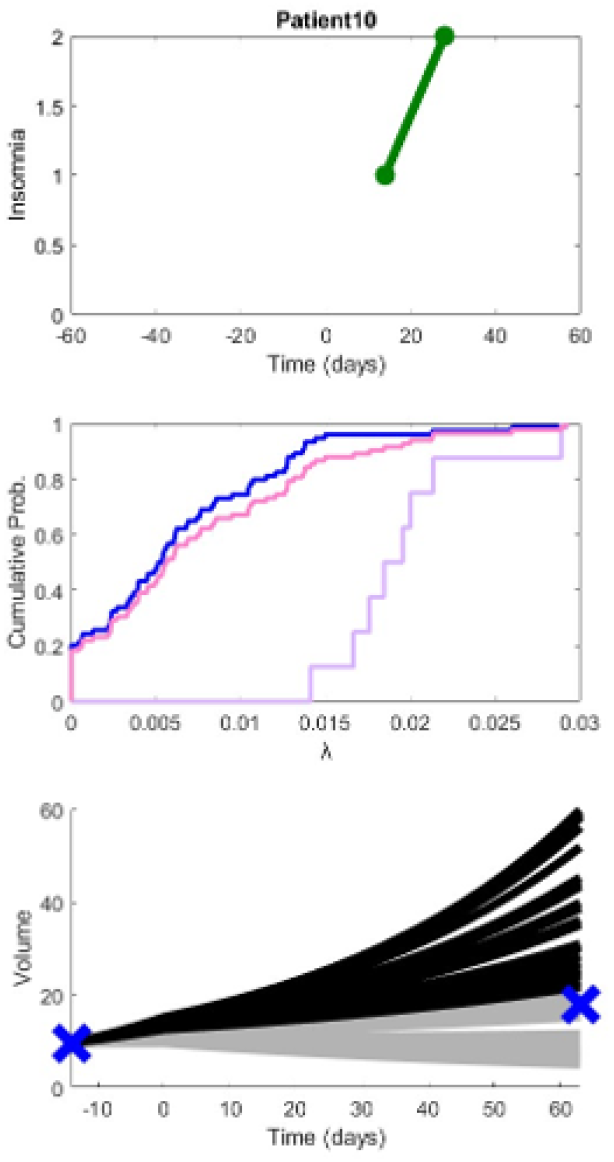
Generating predictions by incorporating insomnia and tumor volume. Patient 10 is correctly predicted as a progressive patient with a resistance rate of 0.28.

The tumor growth inhibition model alone was able to predict treatment response with an overall 66.25% accuracy from volume points one to two. This includes a sensitivity of 75% with six out of eight correct predictions of progression and a specificity of 65.28% with 47 out of 72 correct predictions of stable disease. Similarly, the same analysis was done from volume points one to three and an accuracy of 62.5%. This includes a specificity of 50% with four out of eight correct predictions of progression and a specificity of 64.58% with 31 out of 48 correct predictions of stable disease.

### Incorporating Insomnia Changes Improves Predictability

As seen in the ROC curve of Figure 4, eight progressive patients from one to two volume points rendered eight true positives and two false negatives for a sensitivity of 80%. 72 stable disease patients rendered 85 predictions, which were predicted as 58 true negatives and 27 false positives, rendering a specificity of 68.2%. The sensitivity and specificity for the 80 patients were plotted in the ROC Curve of Figure 4, resulting in a mean area under the curve (mAUC) score of 0.78. The same methodology was reproduced for 55 patients to predict volume dynamics at the third volume measurement. Eight progressive patients from measurements one to three were predicted as 5 true positives and 3 false negatives, rendering a sensitivity of 62.5%. 46 stable disease patients from measurements one to three rendered 54 predictions with 41 true negatives and 13 false positives, for a specificity of 75.9%, generating an mAUC score of 0.60,. Overall, our combined accuracy was 71.3%, and our combined specificity increased to 72.2%.

**Figure 4.**
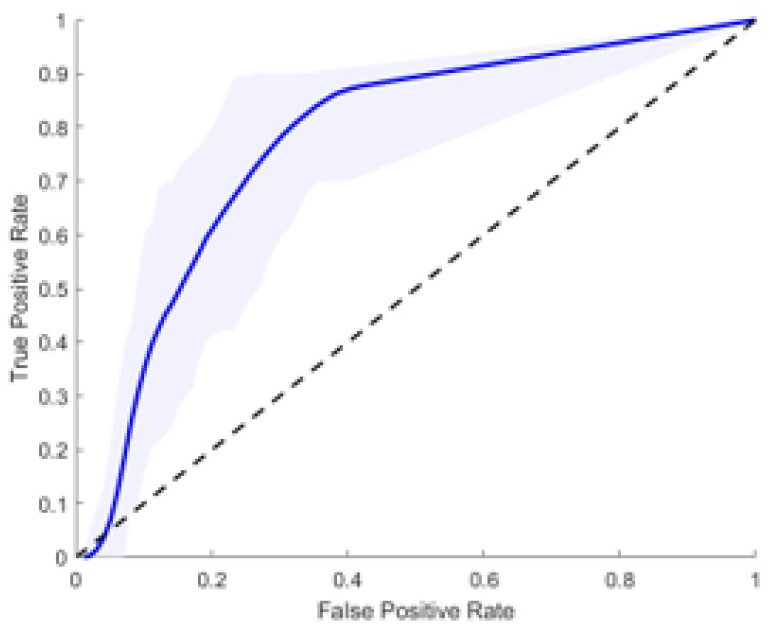
ROC Curve of generated resistance rates after incorporating insomnia data. Sensitivity increased to 72.2%

**Figure 5.**
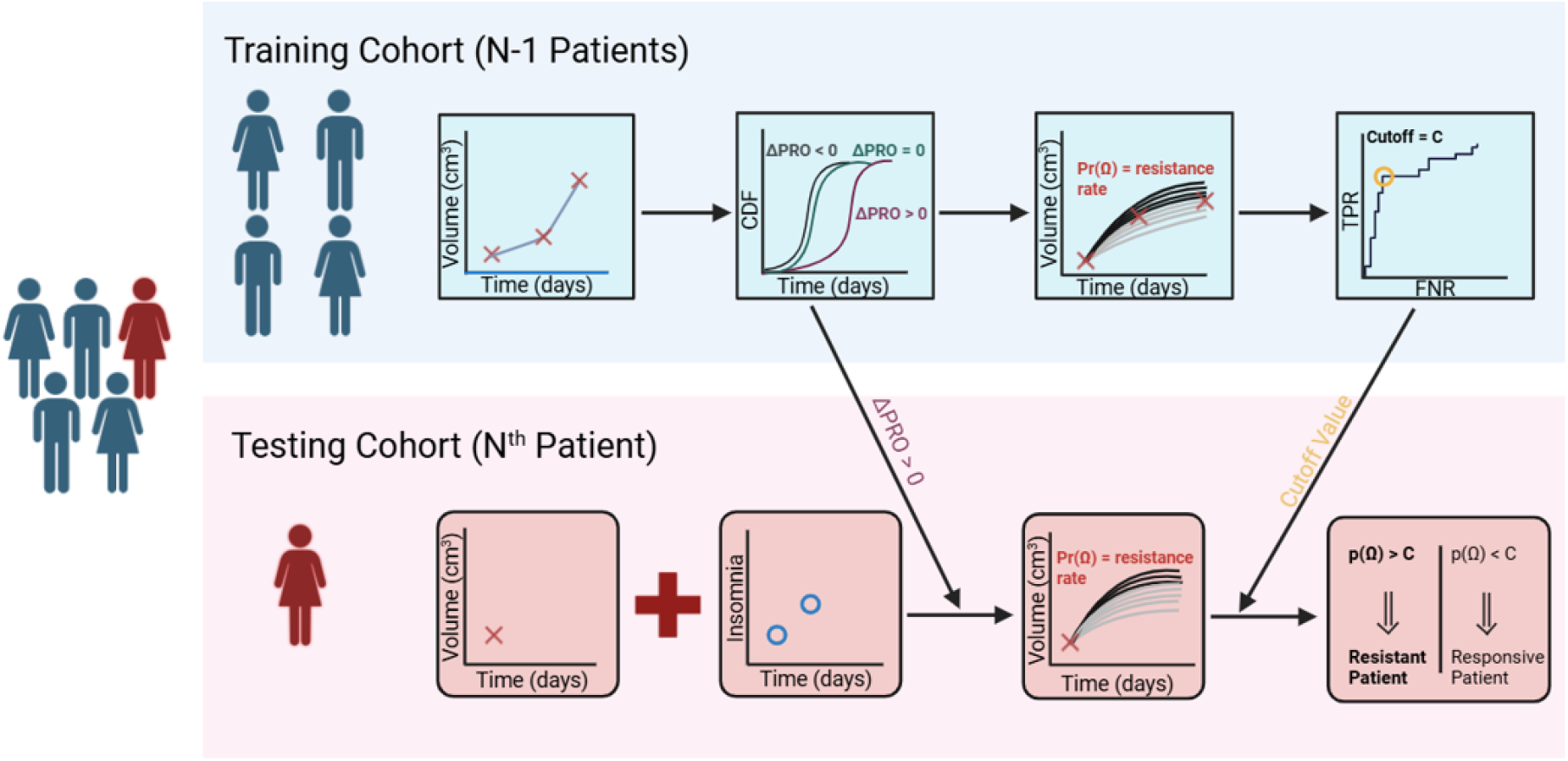
Pipeline of how *N* ^th^ patients are classified as responsive or resistant.

**Figure 6.**
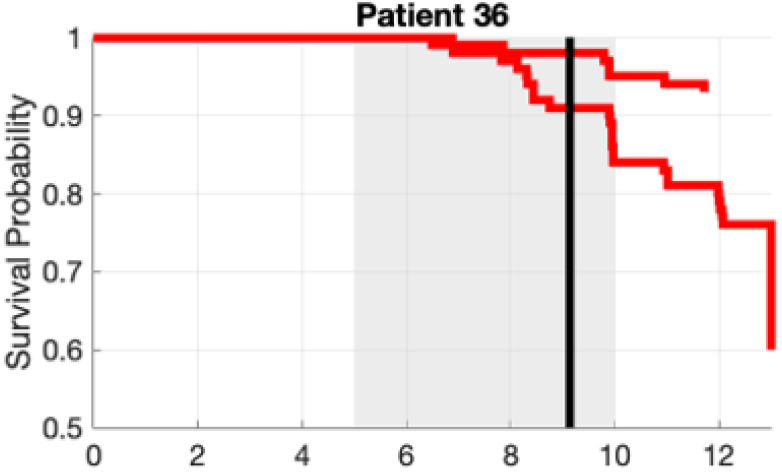
Kaplan Meier plot of progressive patient from first to second volume

#### Addressing the When

We utilized Kaplan-Meier plots to predict precisely when a patient would progress in their disease. Kaplan-Meier curves plot the cumulative distribution of survival against time. Survival corresponds to any “event” of interest. In the context of this study, the event of interest was cancer progression. To calculate survival, serial time, the status of the event at each time, and the study group are inputted (Rich et al. 2014). In our case, the endpoint would correspond to reaching cancer progression. These figures give us insight into the survival probability throughout their treatment on a volume-to-volume basis. Clinicians can therefore identify patients with lower survival probabilities as a possible risk for progressing and intervene.

## Discussion

The goal of this paper was to determine the predictive efficacy of PROs, specifically insomnia, in conjunction with the TGI model. With the amalgamation of these methods, clinicians would be able to predict cancer progression, precisely when it will occur, and consequently intervene to prevent it. Through validation and calibration of the TGI model, we found it to be effective in predicting the behavior of responsive patients with a 94% accuracy, but only 4/13 resistant patients were correctly predicted. Therefore, the inclusion of PROs was necessary to improve the sensitivity. Through a leave-one-out approach, our model incorporated the volume dynamics described by the TGI model and the severity of insomnia changes. This method improved our sensitivity to 72.2%. Although our specificity was lowered to 71.2%, we prioritized the prediction of progression as it has more relevant clinical implications for intervention. The TGI model has proved efficacious at predicting responsive disease, but the incorporation of PROs greatly enhanced our ability to predict resistant disease.

## Limitations

Despite the promising results of the work, we have identified a few limitations to our study, namely, a small, unbalanced dataset. During the cutoff analysis, the disproportionate number of patients in each class made it difficult to choose a threshold. For the second volume measurement predictions, there were 10 progressive events versus 85 responsive events. More frequent PRO surveys and a large sample size could also combat the imbalanced data. Future work could also address the limitations of the model, such as accounting for tumor heterogeneity. As seen in Figure 1, many of the responsive patients hit the lower bound of *λ*, indicating the model is unable to converge for certain cases. Further work could therefore be done on parameter estimation or modifying optimization strategies to account for this.

## Conclusion

All in all, we want to highlight the potential of PROs as minimally invasive biomarkers in tandem with mathematical modeling to predict when progression will occur. We have successfully shown the predictive efficacy of treatment response by incorporating both insomnia as a PRO measure and the TGI model. The TGI model alone, while effective at predicting progression, is unable to accurately predict stable disease. The insomnia data allows us to remedy this problem and obtain an accuracy of 71.3%.

## Data Availability

All data produced in the present study are available upon reasonable request to the authors.

